# Prevalence of drug resistance-conferring mutations associated with isoniazid and rifampicin resistant *Mycobacterium tuberculosis* in Ethiopia: A systematic review and meta-analysis

**DOI:** 10.1101/2020.06.07.20124958

**Authors:** Melese Abate Reta, Birhan Alemnew, Biruk Beletew Abate

**Affiliations:** Department of Medical Laboratory Science, College of Health Sciences, Woldia University, Woldia, Ethiopia; Department of Medical Microbiology, Faculty of Health Sciences, University of Pretoria, South Africa; Department of Nursing, College of Health Sciences, Woldia University, Woldia, Ethiopia

**Keywords:** *M.tuberculosis*, *gene mutations*, antibiotics resistance, *meta-analysis*, *Ethiopia*

## Abstract

**Background:** Drug resistance tuberculosis (DR-TB) continues to be a major public health threat globally. Due to the development of many rapid molecular diagnostic tools to detect gene mutations in *M.tuberculosis (Mtb)*, specific genes conferring resistance to different anti-TB drugs have been identified. The aim of this meta-analysis was to assess the prevalence of the gene mutations associated with rifampicin (RIF) and isoniazid (INH) resistant *Mtb* in Ethiopia.

**Methods:** Using PRISMA guideline, we systematically searched a literature on PubMed/MEDLINE, Web of Science, Scopus electronic databases, Cochrane library, and other database sources. The data analysis was done using STATA 11. The pooled prevalence of the gene mutations associated with resistance to RIF and INH were estimated using the random effect model. Heterogeneity was measured by the I^2^ statistical test, and the publication bias through the funnel plot and the Egger’s regression test.

**Results:** Among all antimycobacterial resistance tested TB patients, prevalence of resistance to any anti-TB drug was 31.3%, while multidrug resistance TB (MDR-TB), any RIF and INH resistance were 22.2%, 24.9%, and 27.9%, respectively. In total, 909 (95.8%) of 949 INH resistant *Mtb* isolates had detectable gene mutation in *kat*G315 and 5.9% in the *inh*A gene. The meta-analysis derived an estimated pooled prevalence of *kat*GMUT1(S315T1) in INH resistant *Mtb* was 89.18% (95%CI 81.94-96.43%), while a pooled *inh*AMUT1 (C15T) resistant *Mtb* prevalence was 77.48% (95% CI 57.84-97.13%). Besides, 769 (90.8%) of 847 RIF resistant strains had detectable *rpo*B gene mutation, commonly in *rpo*BMUT3(S531L) probe (550 cases). The meta-analysis resulting a pooled *rpo*BMUT3(S531L) resistant *Mtb* prevalence of 74.20 % (95%CI 66.39-82.00%).

**Conclusions:** RIF resistant *Mtb* isolates were spread widely, mainly with S531L mutation. Similarly, INH resistant *Mtb* isolates were spread with S315T1 and C15T mutations. It is significant to detect S531L among RIF resistant and S315T1 and C15T mutations among INH resistant isolates as it may be a determinant for subsequent development of MDR-TB. Rapid diagnosis of RIF and INH resistant *Mtb* strains in TB patients would expedite modification of treatment regimens, and proper infection control interventions could be taken on time to reduce the risk of further development and transmission of MDR-TB.

**Highlights:** Tuberculosis(TB), particularly drug resistance TB (DR-TB) continues to be a major public health threat globally. Herein, we used a systematic literature search on reliable electronic databases, and perform a meta-analysis to assess the prevalence of the gene mutations associated with rifampicin (RIF) and isoniazid (INH) resistant Mtb in Ethiopia. The meta-analysis derived an estimated pooled prevalence of *kat*GMUT1 (S315T1) in INH resistant *Mtb* was 89.18% (95%CI 81.94-96.43%), while *inh*AMUT1(C15T) resistant *Mtb* prevalence was 77.48% (95% CI 57.84-97.13%). Besides, among 90.8% of RIF resistant strains which had detectable *rpo*B gene mutation, a pooled S531L resistant *Mtb* prevalence was 74.20% (95%CI 66.39-82.00%). This review revealed that the RIF resistant *Mtb* strains were spread widely, mainly with S531L mutation, while the INH resistant *Mtb* isolates were spread widely with S315T1 and C15T mutations. So, it is significant to detect S531L among RIF resistant and S315T1 and C15T mutations among INH resistant isolates as it may be a determinant for subsequent development of MDR-TB. Rapid diagnosis of RIF and INH resistant *Mtb* strains in TB patients would expedite alteration of treatment regimens, and proper infection control interventions could be taken on time to reduce the risk of further development and transmission of MDR-TB.

## Background

Tuberculosis (TB) disease caused by *Mtb* bacilli, continues as major public health threat globally [1, 2]. Despite the “TB incidence and mortality declined over the past decades, there were still an estimated 10.0 million new cases of TB and approximately 1.45 million deaths attributed to TB worldwide in 2018” [1]. The emergency of antimycobacterial drug resistance is threatening the TB prevention and control activities, and it remains to be a major public health threat on global scale [2]. “In 2018, there were about half a million new cases of RIF resistant TB, of which 78% had MDR-TB” [1].

Anti-TB drug resistance in *Mtb* arises as a result of spontaneous gene mutations that reduce the bacterium resistant to the most commonly used anti-TB drugs. These genes can encode drug targets or drug metabolism mechanisms and influence the effectiveness of anti-TB treatments [1-5]. Inappropriate treatment and patient’s poor adherence to anti-TB drug treatments contribute to the development of drug resistant TB, while the lack of drug resistance diagnosis and subsequent improper TB patients treatment rise the risk of direct transmission of DR-TB [3, 4, 6].

Due to the lack of accurate, rapid, and inexpensive diagnostic tests, there is low drug resistance, including MDR-TB detection rates in resource limited countries. Sputum smear microscopy, the most frequently used diagnostic methods for the “detection of TB disease does not detect drug resistance” [1]. Mycobacterial culture on liquid or solid media, standard drug sensitivity testing (DST), can take longer time to obtain test results, initiate proper anti-TB drug treatment and it needs well-furnished laboratory settings and substantial biosafety resources. This is impracticable in many low resource settings [1, 2, 4, 7]. During the past decade, several molecular (genotypic) DST methods which can detect gene mutations that confer drug resistance have been developed, including line probe assays, real-time polymerase chain reaction (PCR), deoxyribonucleic acid (DNA) sequencing, DNA hybridization on designed chips, and pyrosequencing [3, 8-10]. In pursuance of these molecular diagnostic approaches to correctly identify all resistant *Mtb*, the genes and specific nucleotide change conferring antitubercular drug resistance should be known and included in the diagnostic test. However, “geographical frequency and global distribution of RIF and INH resistance associated *Mtb* gene mutations have not yet been thoroughly measured in the pathogen population” [11].

Several previous review reports have identified different genes that encode anti-TB drug targets and have briefed different mechanisms of resistance to both RIF and INH [12-17]. More than 95% of RIF resistance is associated with the gene mutations in an 81 base pair section of the *rpo*B gene. The INH resistance appears more complex and has been associated with multiple genes, most commonly *kat*G and at the promotor region of *inh*A gen [14, 17-21]. The current molecular diagnostic tests for INH resistance have focused on the detection of the “canonical” mutations in codon 315 of *kat*G and position-15 in the *inh*A promoter region. Many earlier studies have identified highly variable frequencies of these mutations; with *kat*G315 mutations accounting for 42 to 95% and *inh*A-15 mutations accounting for 6 to 43% of phenotypic INH resistance [3, 4, 12, 16, 17, 21-23]. The recent invention of “Xpert MTB/RIF” test [24] and the Line probe assays [25] “which span 81-base pair fragment of the ribonucleic acid (RNA) polymerase beta subunit (*rp*oB) gene have allowed for the rapid detection of resistance to RIF”.

To date there is no systematic review and meta-analysis that has assessed the most common gene mutations conferring RIF and INH resistance in *Mtb* in Ethiopia. Moreover, the pooled estimated prevalence of RIF resistance associated gene mutation, and the frequencies for co-occurring or multiple mutations have not been evaluated in order to understand the overall proportion of phenotypic INH and RIF resistance explained by the existing single or canonical gene mutations.

Hence, it is critically significant to understand the frequency and prevalence of drug resistance-conferring mutation associated with RIF and INH resistant *Mtb in* Ethiopia. “A failure to account for these variations limits the local effectiveness of molecular diagnostic tools currently available and constrains the development of improved genotypic diagnostic tests” [26]. Therefore, the aim of this meta-analysis was to estimate the frequency and prevalence of the most common gene mutations associated with phenotypic RIF and INH drug resistance in *Mtb* in Ethiopia based on the previously published literature’s data.

## Methods

### Study protocol

We strictly followed “the Preferred Reporting Items for Systematic review and Meta-analysis (PRISMA)” guideline [27] to searching records from the databases, paper screening by title, abstract, and evaluation of full text’s eligibility (Fig.1). The completed PRISMA checklist was provided as supplementary file (Table S1). This review protocol have submitted to the International Prospective Register of Systematic Reviews (PROSPERO) on May 2020, and assigned the submission identification number (ID# 186705).

**Figure. 1:**
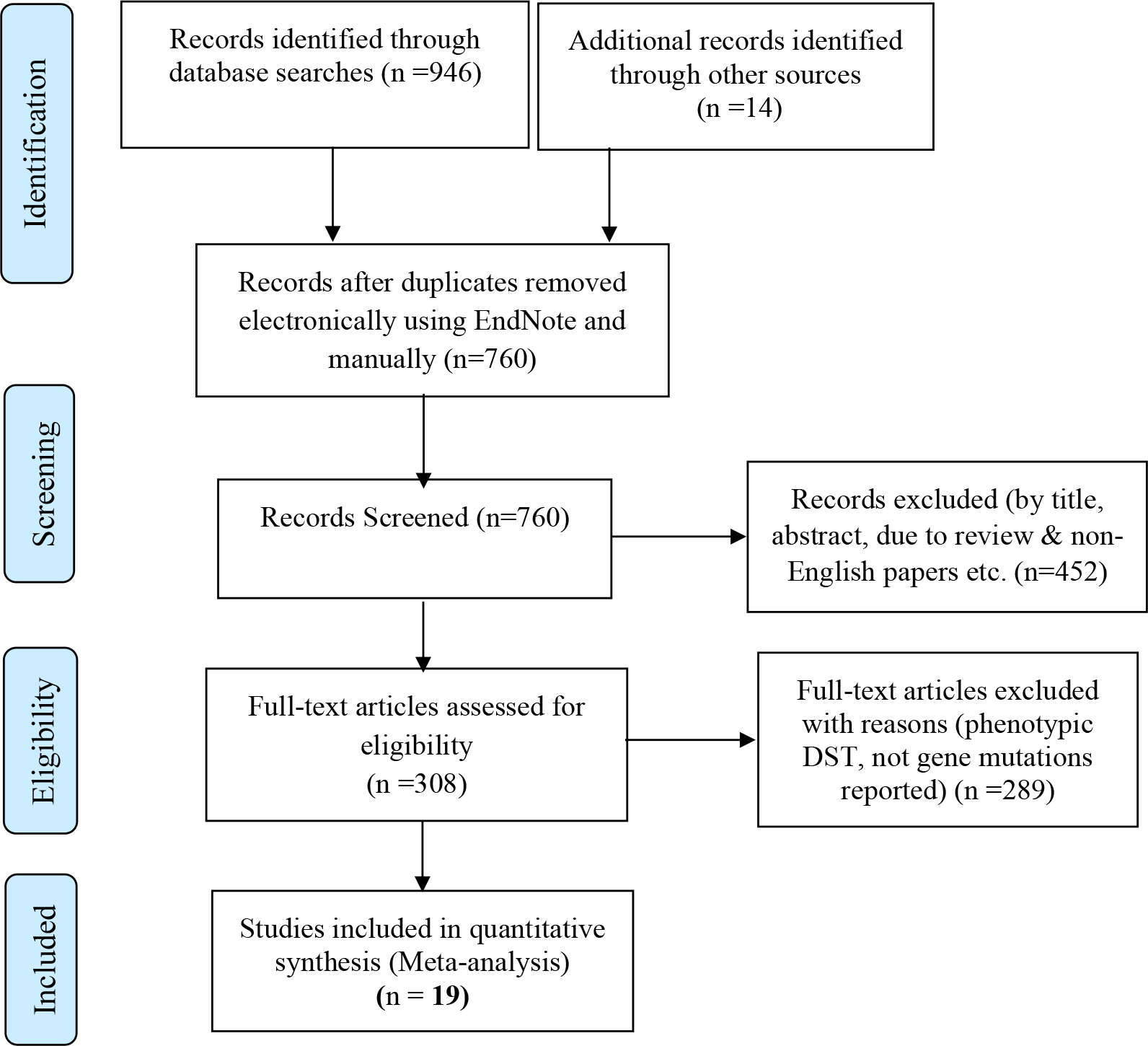
PRISMA –adapted flow diagram showed the results of the search and reasons for exclusion of articles [27]

### Databases and Search Strategy

The research papers published in English language were searched out on PubMed/MEDLINE, Web of Science, and Scopus electronic databases, Cochrane library and other database sources, without restricting the studies publication year. Research studies reported the gene mutations conferring RIF and INH resistance in *Mtb* in Ethiopia were included in the analysis. We used the following specific-subjects headings for databases searching: “*Mycobacterium tuberculosis*”, “tuberculosis”, “drug resistance”, “drug susceptibility testing”, “anti-tuberculosis drug resistance”, “antitubercular agents”, “first-line antitubercular drugs”, “isoniazid resistance tuberculosis”, “rifampicin resistance tuberculosis”, “gene mutations”, “drug resistance-conferring mutations”, “frequency of gene mutations”, “antitubercular drug resistance determinants”, “magnitudes of gene mutations”, “molecular diagnostics”, “molecular detection”, “molecular characterization”, “genotyping”, “line probe assay”, “GenoType®MTBDR*plu*s assay”, “GeneXpertMTB/RIF assay”, “GenoType®MTBDR*sl* assay” and “Ethiopia”. The search strings were applied using “AND” and “OR” Boolean operators. The PubMed key search terms used were: (*Mycobacterium tuberculosis* [MeSH Terms] OR tuberculosis [MeSH Terms]) AND (INH OR isoniazid [MeSH Terms]) AND (RIF OR rifampicin [MeSH Terms]) AND (resistance OR resistant) AND (mutations [MeSH Terms]) OR sequence) AND Ethiopia. The full searching strategy detail for PubMed/MEDLINE database was provided as supplementary file (Table S2). Furthermore, we have reviewed the primary studies and review article’s reference lists to access potential studies and other grey literatures.

### Screening and eligibility of studies

Papers retrieved from these databases were exported into the “EndNote reference software version 8.2 (Thomson Reuters, Stamford, CT, USA)”. Duplicate papers were sorted, noted and removed using the EndNote software. Some duplicated papers were identified manually due to the presence of difference in reference writing formats across the databases. Subsequently, two researchers (MAR and BA) independently evaluated the paper’s title and abstracts using the preset inclusion criteria. Two investigators (BA and BBA) also have independently collected full texts and assessed the eligibility of articles for final inclusion to the analysis. In each case, the discrepancies arose between two authors were resolved through discussion with the other authors to come into consensus.

### Inclusion and Exclusion Criteria

All observational studies (cross-sectional, case-control, and cohort) which have diagnosed RIF and INH resistance in *Mtb* using standard World Health Organization (WHO) approved molecular DST tools, and had reported mechanisms of anti-tuberculosis drug resistance / or the gene mutations conferring RIF and INH resistance in *Mtb* in Ethiopia were included. Furthermore, studies addressing frequencies of the gene mutation, and some or all of the following criteria were included: (a) Studies reported data concerning the prevalence of antitubercular drug resistance among pulmonary and extra pulmonary TB patients (both retreated or newly diagnosed cases); (b) The prevalence of any ant-TB drug resistance or MDR-TB or extensive drug resistance TB (XDR-TB); (c) Studies which used standard WHO approved molecular anti-TB drug susceptibility diagnostic methods; (d) TB research conducted in Ethiopia and published in English language. We excluded those studies from the analysis with the following exclusion criteria: (a) the studies which were not reported mechanisms of ant-tuberculosis drug resistance/ or the gene mutations conferring RIF and INH resistance in *Mtb*; (b) The studies reported data on non-tuberculous mycobacterium; (c) Studies which did not assess DST of RIF and INH; (d) Studies performed anti-TB DST only through phenotypic methods. The editorial’s report, narrative review studies, case reports, and qualitative studies were excluded from the analysis. Additionally, we excluded citations without full text after contacting a study author twice through email.

### Quality assessment

The critical quality assessment checklist recommended by the “Joanna Briggs Institute (JBI) was used to evaluate the quality of all included studies” [28]. The three investigators (MAR, BA and BBA) independently evaluate the quality of the full text articles. The discrepancy was resolved through discussion to reach on consciences, and to include articles to final analysis. The domain paper quality assessment criteria were; clear inclusion criteria, details of study subjects and the study settings, reliable/valid measurements for exposure, outcome variables and appropriate statistical analysis. Studies (case-control, cross-sectional and cohort) with the score of four and above were considered good quality and included, while the studies have the average score of three and below were considered as poor quality, and excluded (Table S3).

### Data extraction

We used standard data extraction format prepared in Microsoft Excel sheet. Two authors (MAR and BA) independently extracted the following relevant information related to study characteristics (Autor/s’ name, publication year, study period, study region, type of TB patients, study design, molecular DST method/s, sample size, total positive cases, total *Mtb* isolates which DST was performed, frequency of any anti-TB drug resistance, any INH and RIF resistance and MDR-TB, and RIF and INH resistance mechanisms/frequency of gene mutations (*rpo*B, *kat*G and *inh*A) and frequency of specific amino acid/nucleotide (codon) changes in each resistant gene loci/probe; at ***rpo*B** gene [*rpo*BMUT1 (D516V), *rpo*BMUT2A (H526Y), *rpo*BMUT2B (H526D), and *rpo*BMUT3 (S531L)], at ***kat*G** gene [*kat*GMUT1 (S315T1) and *ka*tGMUT2 (S315T2)], at ***inh*A** [*inh*AMUT1 (C15T), *inh*AMUT2 (A16G), *inh*AMUT3A (T8C), and *inh*AMUT3B (T8A)]. Furthermore, the absence of wild-type probe at each gene loci (*rpo*BWT1-8, *kat*GWT1 and *inh*AWT1&2) were evaluated (Table 1 and Table S4).

**Table 1.** Characteristics of included studies.

| Author/s | Publication year | Study period | Study region | Type of patients | Study design | Molecular diagnostic methods | Number of patient (participants) (n) | Total positive cases | Total isolates which DST is performed(n) | Any drug Resistance, (n) | Any RIF and INH resistance, (n) |  | MDR, TB (n) | Anti-tuberculosis drug resistance mechanisms (n) | Frequency of gene mutations (n) |  |  |  |  |
| --- | --- | --- | --- | --- | --- | --- | --- | --- | --- | --- | --- | --- | --- | --- | --- | --- | --- | --- | --- |
|  |  |  |  |  |  |  |  |  |  |  | INH, (n) | RIF, (n) |  |  | rpoB | katG | inhA | katG+inhA | rpoB+katG |
| Abate et al [30] | 2014 | 2012 - 2013 | AA | PTB | Retrospective cross-sectional | GenoType®MTB DR<sub>plus</sub> assay | 736 | 736 | 736 | 523 | 481 | 470 | 427 | rpoB: S531 L(323), H526Y (31), H526D (19), D516V (13), rpoBWT2 (3), rpoBWT3 (8), rpoBWT7 (28), rpoBWT4 (21), rpoBWT6 (2), rpoBWT8 (12), rpoBmixed mutations at different codon positions (10); katG: S315T1 (447), S315T2 (5), katGWT mutation (315)-unknown codon (9), katG mixed mutations (S315T) (4); inhA: C15T (5), inhAWT2mutation (1), katGMUT1 + inhAMUT1 (S315T+C15T) (9), katGMUT1 + inhAMUT2 (S315T+A16G) (1). | 470 | 469 | 6 | 10 | 0 |
| Alelign et al [31] | 2019 | 2015-2017 | AM | PTB & EPTB | Cross-sectional | GenoType MTBDR<sub>plus</sub> assay | 111 | 111 | 111 | 20 | 20 | 2 | 2 | rpoB: rpoBWT6 (2), rpoBWT8 (2); katG: S315T1 (20), katGWT (6); inhAWT1 (20), inhAWT2 (20) | 2 | 20 | 20 | 20 | 20 |
| Bedewi et al [32] | 2016 | 2012 - 2013 | OR | PTB | Cross-sectional | GenoType®MTB DR<sub>plus</sub> assay | 279 | 279 | 279 | 31 | 25 | 9 | 3 | rpoB: S531L (3), H526Y (2), rpoBWT8 missed (4); katG: S315T1 (22), katGWT (3); inhA: C15T (2), inhAWT2 missed (1). | 9 | 25 | 3 | NR | NR |
| Bekele et al [33] | 2018 | 2006-2010 | AA, AM, OR, SNNP R | PTB & TBLN | Cross sectional | GenoType®MTB DR<sub>plus</sub> assay | 950 | 161 | 161 | 14 | 12 | 7 | 5 | rpoB: D516V (8), S531L (3); H526Y (7), H526D (7), rpoBW8 missed (4); katG: S315T2 (8), S315T1 (3), katGWT missed (8); inhA: C15T (6), A16G (7), T8C (8), T8A (7), inhAWT1 (2). | 8 | 8 | 8 | NR | NR |
| Beyene et al [34] | 2009 | 2005-2006 | SNNP | EPTB | Cross-sectional | MLPA | 171 | 156 | 95 | 11 | 11 | NR | NR | KatG: S315T (11). | NR | 11 | N R | NR | NR |
| Biadlegne et al [35] | 2014 | NR | AM | EPTB | Cross-sectional | GeneXpertMTB/ RIF assay & GenoType®MTB DR<sub>plus</sub> | 231 | 32 | 32 | 3 | NS | 3 | 3 | rpoB: S531L (2), #rpoBWT3 (deletion) (1). | 3 | NR | NS | NS | NR |
| Biadlegne et al [36] | 2013 | 2012 | AM | EPTB | Cross-sectional | GenoType®MTB DR<sub>plus</sub> & GenoType® MTBDRs/ | 226 | 226 | 226 | 13 | 8 | 3 | 2 | rpoB: S531L (2), Q513L (1), rpoBWT3(1), rpoBWT8(2); katG: S315T (8), katGWT(8). | 3 | 8 | 0 | NR | 2 |
| Brhane et al [37] | 2017 | NR | SO | PTB | Cross-sectional | GenoType®MTB DR<sub>plus</sub> | 105 | 98 | 98 | 18 | 18 | 10 | 10 | rpoB: S531L (8), H526TY (1), H526D (1); katG: S315T1 (15), katGWT (15); inhA: C15T (3). | 8 | 15 | 3 | NR | NR |
| Damena et al [38] | 2019 | 2015-2016 | AA | PTB | Cross-sectional | GenoType®MTB DRplus & GenoType MTBDRs/ | 213 | 150 | 150 | 20 | 20 | 16 | 16 | *rpoB*: S531L (13), D516V (1), H516Y (1), H516D (3), *rpoBWT87* (13), *rpoBWT7* (2); *KatG*: S315T1 (20), *katGWT* (19). | 15 | 20 | 0 | 0 | 15 |
| Haile et al.[39] | 2020 | 2015-2016 | OR | PTB |  | GenoType MTBDRplus | 111 | 92 | 92 | 6 | 5 | 1 | 0 | *rpoB*: S531L(1), *rpoBWT8* (1); *katG*: S315T1 (5), *katGWT* (5). | 1 | 5 | 0 | 0 | 0 |
| Kebede et al [40] | 2017 | 2011/12 | AA | MDR-TB | NR | GenoType®MTB DRplus | 72 | 72 | 72 | 53 | 52 | 49 | 48 | *rpoB*: S531L (40), D516V (2), H526Y (4), *rpoBWT8* (35), *rpoBWT2* (1), *rpoBWT4* (2), *rpoBWT3* (4), *rpoBWT7* (6); *katG*: S315T1 (52). | 46 | 52 | N R | NR | NR |
| Meaza et al [41] | 2017 | 2015 | AA | MDR-TB | Cross sectional | GenoType®MTB DRplus V 2.0 assay | 274 | 89 | 89 | 49 | 37 | 41 | 29 | *rpoB*: S531L (27), H526Y (2), *rpoBWT8* (27), *rpoBWT7* (2), *rpoBWT1* (1); *katG*: S315T/L (28); *inhA*: C15T (1), A16G (1), *inhAWT1* (1), *inhAWT2* (1). | 27 | 28 | 1 | 1 | NR |
| Mekonnen et al [42] | 2015 | 2012-2014 | AM | MDR-TB | Cross-sectional | GenoType®MTB DRplus assay | 413 | 413 | 413 | 195 | 176 | 169 | 150 | *rpoB*: S531L (85), H526Y (19), H526D (7), *rpoBWT2* (13), *rpoBWT3* (33), *rpoBWT4* (23), *rpoBWT7* (39), *rpoBWT8* (95); *katG*: S315T1 (155), Dele-315 (16), S315T2 (1), *katGWT* (172); *inhA*: C15T (8), *inhAWT1* (8). | 111 | 155 | 8 | 3 | NR |
| Tadesse et al [43] | 2017 | 2013-2015 | OR | EPTB | NR | GeneXpertMTB/ RIF assay | 436 | 310 | 279 | 10 | NS | 10 | 10 | *rpoB*: (codon 447-452) (10) | 10 | NS | N S | NS | NS |
| Tadesse et al [44] | 2016 | 2013-2014 | OR | PTB | Cross-sectional | GenoType®MTB DRplus V.2 assay | 122 | 118 | 112 | 44 | 41 | 34 | 31 | *rpoB*: S531L (28), H526D (1), *rpoBWT7* (2), *rpoBWT8* (31); *katG*: S315T1 (36), *katGWT*-315 (31) (31); *inhA*: C15T (4), *inhAWT* (15/16)(4) | 34 | 36 | 4 | NR | NR |
| Tessema et al [45] | 2012 | 2009 | AM | PTB | Cross sectional | GenoType®MTB DRplus & GenoType®MTB DRs/ | 260 | 260 | 260 | 45 | 35 | 15 | 13 | *rpoB*: S531L (11), H526D (1), *rpoBWT8* (11), *rpoBWT7* (1); *katG*: S315T (33), *katGWT* (33); *inhA*: C15T (2), *inhAWT1* (2). | 15 | 33 | 2 | NR | NR |
| Wondale et al [46] | 2018 | 2014-2016 | SNNP | PTB & EPTB | NR | GenoType®MTB DRplus V. 2.0. | 161 | 126 | 126 | 4 | 1 | 3 | 1 | *rpoB*: H526D (1), D516V(1); *katG*: S315T2 (1) | 3 | 1 | N R | NR | 2 |
| Workalemahu et al [47] | 2013 | 2011 | OR | PTB | NR | GenoType®MTB Rplus assay | 121 | 15 | 15 | 1 | 1 | NR | 0 | *inhA* (1)# | NR | NR | 1 | NR | NR |
| Zewdie et al [48] | 2018 | 2014 | AA | EPTB | Cross-sectional | GenoType®MTB DRplus assay | 65 | 60 | 60 | 6 | 6 | 5 | 5 | *rpoB*: S531L (4), H526Y (1), *rpoBWT8* (4), *rpoBWT7* (1); *katG*: S315T2 (1), S315T1 (5), *katGWT* (6) | 5 | 6 | 0 | 0 | 5 |

**NR**: #= Amino acid change not reported; \*=Frequency of codon changes not reported; AA=Addis Ababa, AM=Amhara, OR=Oromia, HR=Harari, Tig=Tigray, GA=Gambella, AF=Afar, SNNP=South Nation, Nationality and People, SO=Somalia; PTB= Pulmonary tuberculosis, EPTB=extrapulmonary tuberculosis, MDR-TB =multidrug resistance tuberculosis, TBLN=tuberculosis lymphadenitis ; NR=note reported; NS=Not studied; WT=wide type; Mut=Mutant.

### Outcome of interest

This systematic review and meta-analysis had estimated the pooled prevalence of gene mutations conferring RIF and INH resistance in *Mtb* in Ethiopia. Frequency of any anti-tuberculosis drug resistance, and resistance to any INH and RIF were drawn from each included study. The frequency of each resistant gene mutations were counted out of the total resistant *Mtb* isolates for a particular anti-TB drugs. Similarly, the rate of each amino acid/ or nucleotide (codon) changes at each resistant gene locus/probe (*rpo*B, *kat*G and *inh*A) was calculated out of the total resistant gene. The pooled estimate of the prevalence of amino acid/ or nucleotide (codon) changes at each resistant gene loci/probe: ***rpo*B** gene [*rpo*BMUT1 (D516V), *rpo*BMUT2A (H526Y), *rpo*BMUT2B (H526D) and *rpo*BMUT3 (S531L)], at ***kat*G** gene [*kat*GMUT1 (S315T1) and *ka*tGMUT2 (S315T2)], at ***inh*A** [*inh*AMUT1 (C15T), *inh*AMUT2 (A16G), *inh*AMUT3A (T8C) and *inh*AMUT3B (T8A)] were measured. We have also estimated the pooled prevalence of gene mutations/the absence of band at each wild-type probe of ***rpo*B** gene (*rpo*BWT1-8), ***kat*G** gene (*kat*GWT), ***inh*A** gene (*inh*AWT1 and *inh*AWT2)(Table S4).

### Data processing and statistical analysis

The relevant data were extracted from the included studies using standard format prepared in Microsoft Excel sheet and the data were exported into STATA 11.0 for further analysis. Using the binomial distribution formula, Standard error was calculated for each study. “Considering variation in true effect sizes across population, Der Simonian and Laird’s random effects model was performed for the analyses at 95% confidence level”. The heterogeneity of studies was determined using Cochrane’s Q statistics (Chi-square), invers variance (I^2^) and p-values. Publication bias across the studies was measured through the Egger’s regression test [29], and displayed with funnel plots of standard error of Logit event rate. A *p*-value less than 0.05 was considered statistically significant.

## Results

### Search results

As illustrated in figure 1, a total of 960 potential research studies were documented from searched electronic databases and other data sources. Of the total, 760 articles were non-duplicated and subjected for further evaluation; 452 were evaluated and excluded based on their title and abstract, while 308 papers were retained for full-text article review. After full-text article evaluation, 19 studies on the prevalence of gene mutations associated with RIF and INH resistant *Mtb* in Ethiopia were used for final analysis (meta-analysis).

### Characteristics of included studies

As described in Table 1 and Table S4, a total of 19 studies with 5,057 TB patients (3504 culture, line probe assay and/or GeneXpertMTB/RIF assay positive *Mtb* isolates) were included for final analysis [30-48]. Five studies were from Addis Ababa [30, 38, 40, 41, 48], Amhara region [31, 35, 36, 42, 45], and Oromia region [32, 39, 43, 44, 47], respectively, while two studies were from South Nation Nationality and People [34, 46], and one studies from Somalia region [37]. In study design, fourteen studies were cross-sectional [30-38, 41, 42, 44, 45, 48], while five studies [39, 40, 43, 46, 47] have not stated the study design. Nineteen studies, of which eight studies [30, 32, 37-39, 44, 45, 47] have assessed RIF and INH resistance rate among pulmonary TB (PTB) patients, five studies [34-36, 43, 48] have assessed among extra pulmonary TB (EPTB) patients, while three studies [31, 33, 46] have done on both EPTB and PTB patients. GenoType®MTBDR*plus* assay and GeneXpertMTB/RIF assay were the most common molecular

/genotypic DST methods used [30-33, 35-48]. The resistance rate of *Mtb* to any ant-TB drugs, MDR-TB, and resistance to RIF and INH was calculated out of a total of 3406 *Mtb* isolates which their DST was performed [30-48]. In total, 17 studies evaluated the prevalence of any INH resistance [30-34, 36-42, 44-48], and any RIF resistance [30-33, 35-46, 48] among 3406 TB patients. Almost all included studies [30-33, 35-48], except one study had reported the prevalence of MDR-TB strains. All included studies [30-46, 48] had reported the gene mutations associated with resistance to RIF and INH. Moreover, seventeen studies [30-33, 35-46, 48] had quantified the frequency of *rpo*B gene mutation and nucleotide (codon) changes in an 81-base pair *β*-subunit (*rpo*B) gene among 847 RIF resistant *Mtb* isolates, while sixteen studies [30-34, 36-42, 44-46, 48] had reported the frequency of *kat*G gene mutation and nucleotide (codon) changes among 949 INH resistant *Mtb* isolates. Only ten studies [30-33, 37, 41, 42, 44, 45, 47] had reported the gene mutation in *inh*A promotor region, while four studies [30, 31, 41, 42] reported the co-occurrence of *inh*A and *kat*G genes among INH resistant *Mtb* isolates.

### Prevalence of any RIF and INH resistant *M*.***tuberculosis***

Overall, 5057 pooled TB suspected patients were tested by Line probe assays (GenoType^®^MTBDR*plus* v.2.0 and/or GenoType^®^MTBDR*sl assays*) and GeneXpertMTB/RIF assay to identify MDR-TB, RIF and INH resistance pattern [30-48]. Prevalence of any anti-TB drug resistance among all diagnosed TB patients was 31.3% (1066/3406), while the prevalence of any RIF and INH resistant *Mtb* were 24.9% (847/3406), and 27.9% (949/3406), respectively. Moreover, prevalence of MDR-TB was 22.2% (755/3406) (Fig.2 & Table S5). The prevalence of any anti-TB drug resistance rate varies across the studies and geographical locations of Ethiopia. From the included studies, seven studies had reported higher prevalence of any anti-TB drug resistance ranged from 18.0% to 73.6% [30, 31, 37, 40-42, 44]. The prevalence of any INH resistant *Mtb* ranged from 0.8% to 72.2*%*, while the prevalence of any RIF resistant *Mtb* ranged from 1.3% to 68.1% [30-33, 35-46, 48] (Table 1 and Table S5).

**Figure. 2:**
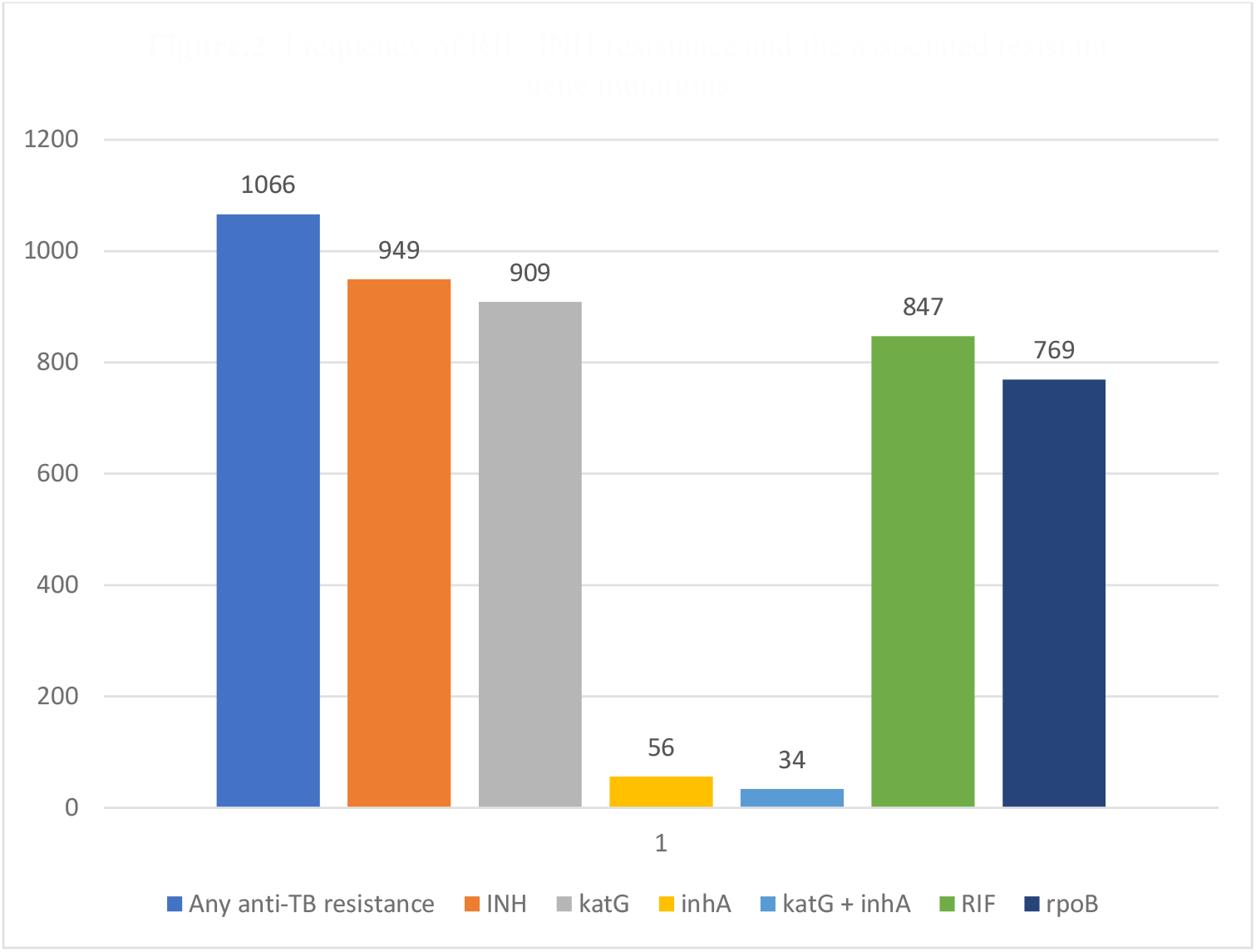
Frequency of RIF, INH resistance and the associated resistant gene mutations

### The frequency of *rpo*B, *kat*G, and *inh*A gene mutations

“The Line probe assay (GenoType^®^MTBDR*plus* assay) (Hain Life science GmbH, Nehren, Germany) strip contains seventeen probes, with amplification and hybridization controls to confirm the diagnostic procedure” [7]. To detect INH resistance, one probe covers the wild-type S315 region of *kat*G gene, while the other two probes (*kat*MUT1 and *kat*GMUT2) are designed to evaluate the S315T (Ser→ Thr) nucleotide (codon) change/ or mutations. Moreover, at the *inh*A promoter region is included on the new strip and covers the regions from positions 15 to 16 for the *inh*AWT1 probe and position 8 for the *inh*AWT2 probe. In *inh*A, four mutations (C15T, A16G, T8C and T8A) can be targeted with the *inh*AMUT1, i*nh*AMUT2, *inh*AMUT3A and *inh*AMUT3B probes. To detect RIF resistance, one probe covers the wild-type probes *rpo*BWT1 to *rpo*BWT8 (507-533) regions of *rpo*B gene, while the other four probes (*rpo*BMUT1, *rpo*BMUT2A, *rpo*BMUT2B, *rpo*BMUT3) are designed to identify the (D516V, H526Y, H526D, and S531L) nucleotide (codon) change/or mutations, respectively. The resistant *Mtb* strain is confirmed when there is absence of one or more wild-type band or probe/s or the presence/ staining of mutant probes (Fig. S1).

A total of 949 any INH resistant *Mtb* strains were identified by standard WHO approved molecular diagnostic methods, among which a higher proportion of mutation was detected in the *kat*G gene (95.8%; 909/949) compared with the gene mutation in the *inh*A promoter region (5.9%; 56/949). In INH resistant *Mtb* strains, the most common mutations were observed in *kat*GMUT1 probe (860 cases) and *kat*WT probe (309 cases). In the *inh*A promoter region, the most frequent mutations were observed in *inh*AMUT1 probe (*inh*A C15T; 31 cases), *inh*AWT1 probe (15/16; 30 cases) and *inh*AWT2 probe (8; 23 cases), while the frequency of mutation in *inh*AMUT2 probe was (15/16; 10 cases). The frequency of mutations in the *inh*AMUT3A and MUT3B were (8; 8 cases), respectively. In this systematic review, a total of 34 *Mtb* strain*s* had mutations in both *kat*G and *inh*A promoter region. The other most frequently occurring mutations in INH resistant *Mtb* isolates, at the position 15 of the *inh*A promotor region, was identified in 3.3% (31/949) of phenotypically resistant isolates, and the mutation in the *inh*A promotor region at a position 8 was identified among 8 (0.8%) of phenotypically resistant strains (Table 1 and Table S5).

Besides, a total of 847 any RIF resistant *Mtb* strains were identified by either Line probe assay or GeneXpert^®^MTB/RIF assay, among which frequency of mutation in the *rpo*B gene was (90.8%; 769/847). In RIF resistant *Mtb* strains, the most common mutations were found in *rpo*BMUT3 (S531L) probe (550 cases), *rpo*BWT8 probe (224 cases), and *rpo*BWT7 probe (91 cases), while the other gene mutations were observed in *rpo*BMUT2A(H526Y) (68 cases), *rpo*BMUT2B (H526D)(40 cases), and *rpo*BMUT1 (D516V) (25 cases). Moreover, the gene mutations at the *rpo*BWT3 and *rpo*BWT4 were 43 and 46 cases, respectively. Besides, ten RIF resistant *Mtb* strains revealed that the *rpo*B gene mutation at (codon 447-452), while the other one strain had *rpo*B gene mutation at CAA/G→ UUA/G(Q513L) (Table 1 and Table S5).

### Meta-analysis

Nineteen studies, of which seventeen studies have evaluated 949 genotypically resistant *Mtb* isolates for mutations in *kat*G gene inclusive of codon 315. In INH resistance, the increasing frequencies of co-occurring mutations were evaluated first by specific genes. This meta-analysis resulting a pooled *kat*GMUT1 (S315T1) resistant *Mtb* prevalence of 89.18% (95% CI 81.94-96.43%) with a I^2^-value of 76.2% and p=0.002 (Fig.3 & Table 2). During our evaluation of publication bias, a funnel plot showed a symmetrical distribution, and the Egger’s regression test p-value was 0.071, which indicated the absence of publication bias (Fig. 4). However, this review derived low pooled prevalence of *kat*GMUT2 (S315T2) resulting 0.91% (95% CI 0.195-1.63%) with I^2^-value of 0.0% and p=0.466 (Table 2). Besides, the pooled estimated prevalence of the absence of band at the wild-type (*kat*GWT(315)) was 48.69% (95% CI -5.20-102.58%) with I^2^-value of 99.5% and p<0.001 (Table 2).

**Figure 3:**
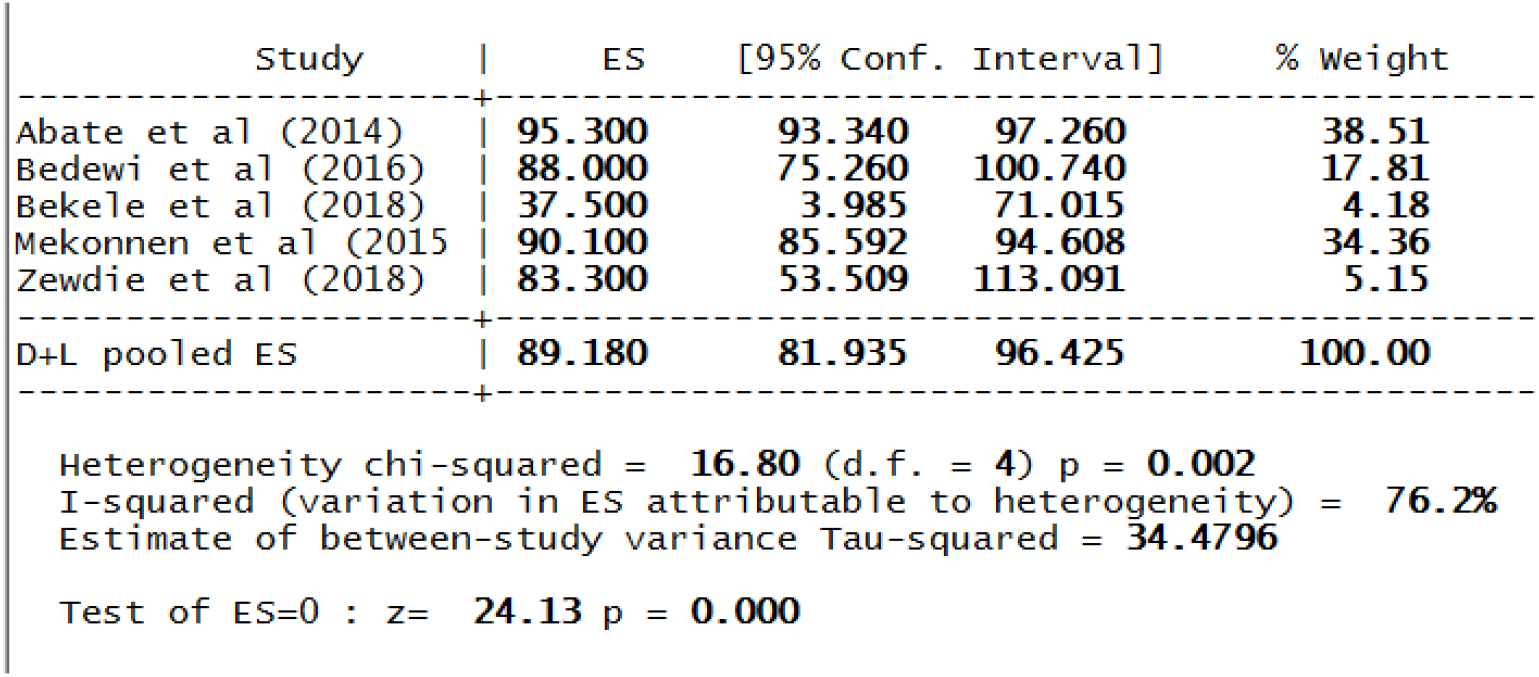
The pooled prevalence of *kat*GMUT1 (S315T1) resistance among INH-resistant *Mtb* cases.

**Figure 4:**
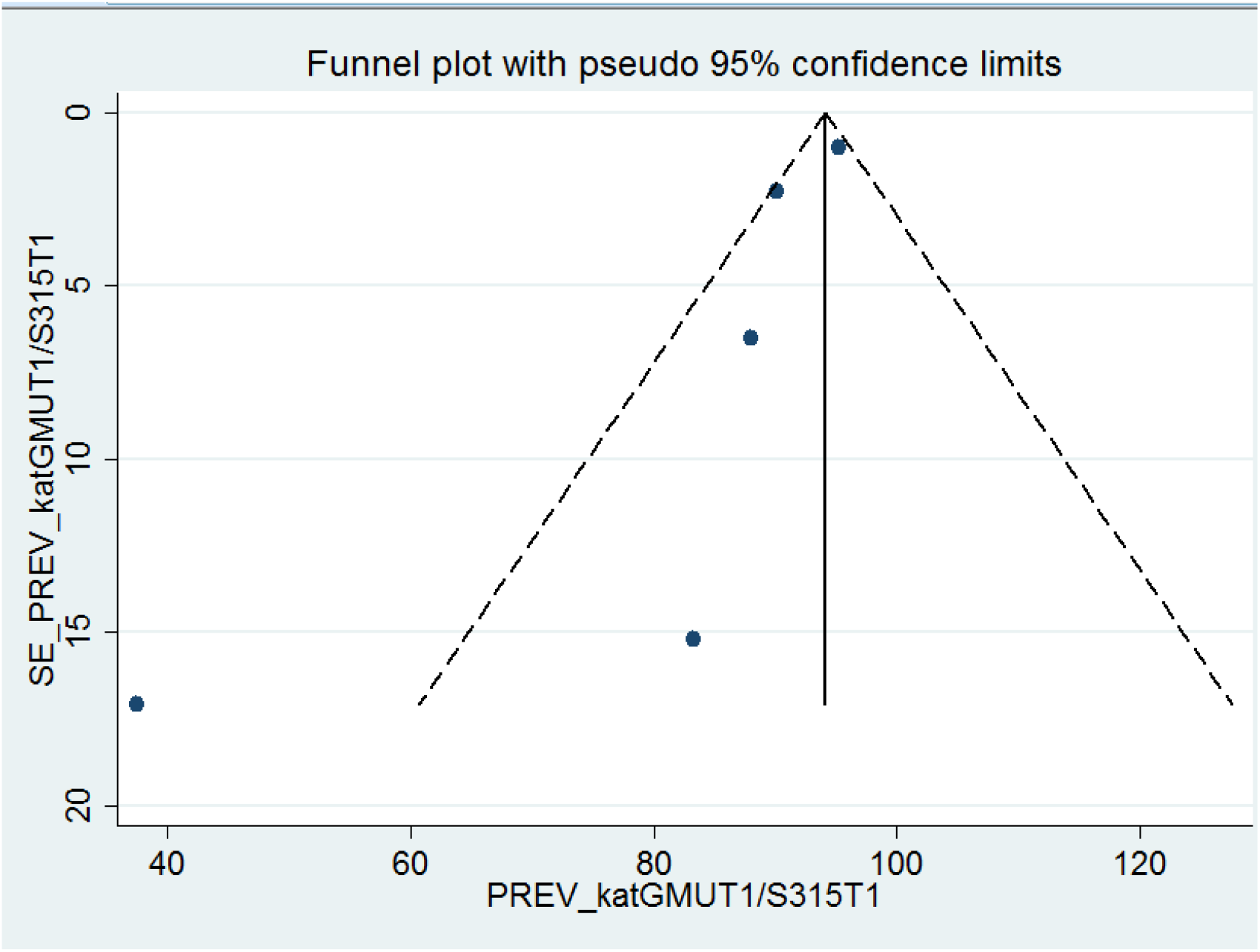
Funnel plot for publication bias, PREV (prevalence) represented in the x-axis and standard error (SE) of prevalence of *kat*GMUT1 (S315T) in the y-axis

**Table 2:** Results of mutations pattern of 949 any INH resistance *tuberculosis* patients.

| Mutation(s) |  | Frequency |  | I<sup>2</sup>(P-value) |
| --- | --- | --- | --- | --- |
| *katG* | *inhA* | No of patients | % (95% CI) |  |
| *katG*WT(315) absent | - | 567 | 48.69 (-5.20-102.58) | 99.5% (<0.001) |
| *katG*MUT1(S315T1) | - | 700 | 89.18 (81.94-96.43) | 76.2% (0.002) |
| *katG*MUT1(S315T2) | - | 663 | 0.91 (0.195-1.63) | 0.0% (0.466) |
| - | *inhA*WT1(15/16) absent | - | - | - |
| - | *inhA*WT2(8) absent | 506 | 20.65 (-5.36-46.66) | 0.0% (0.594) |
| - | *inhA*MUT1(C15T) | 518 | 77.48 (57.84-97.13) | 0.0% (0.848) |
| - | *inhA*MUT2(A16G) | - | - | - |
| - | *inhA*MUT3A(T8C) | - | - | - |
| - | *inhA*MUT3B(T8A) | - | - | - |

The gene mutations at *inh*A promoter region were estimated, and the meta-analysis analysis derived a pooled *inh*AMUT1(C15T) resistant *Mtb* prevalence of 77.48% (95% CI 57.84-97.13%) with I^2^-value of 0.0% and p=0.848 (Fig.5 & Table 2). The publication bias was evaluated using the Egger’s regression test, and p-value was 0.460, which indicated the absence of publication bias. Similarly, the pooled estimated prevalence of the absence of band at wild-type *inh*AWT2(8) resistant *Mtb* was 20.65% (95%CI -5.36-46-66%) with I^2^-value of 0.0% and p= 0.594 (Table 2).

**Figure 5:**
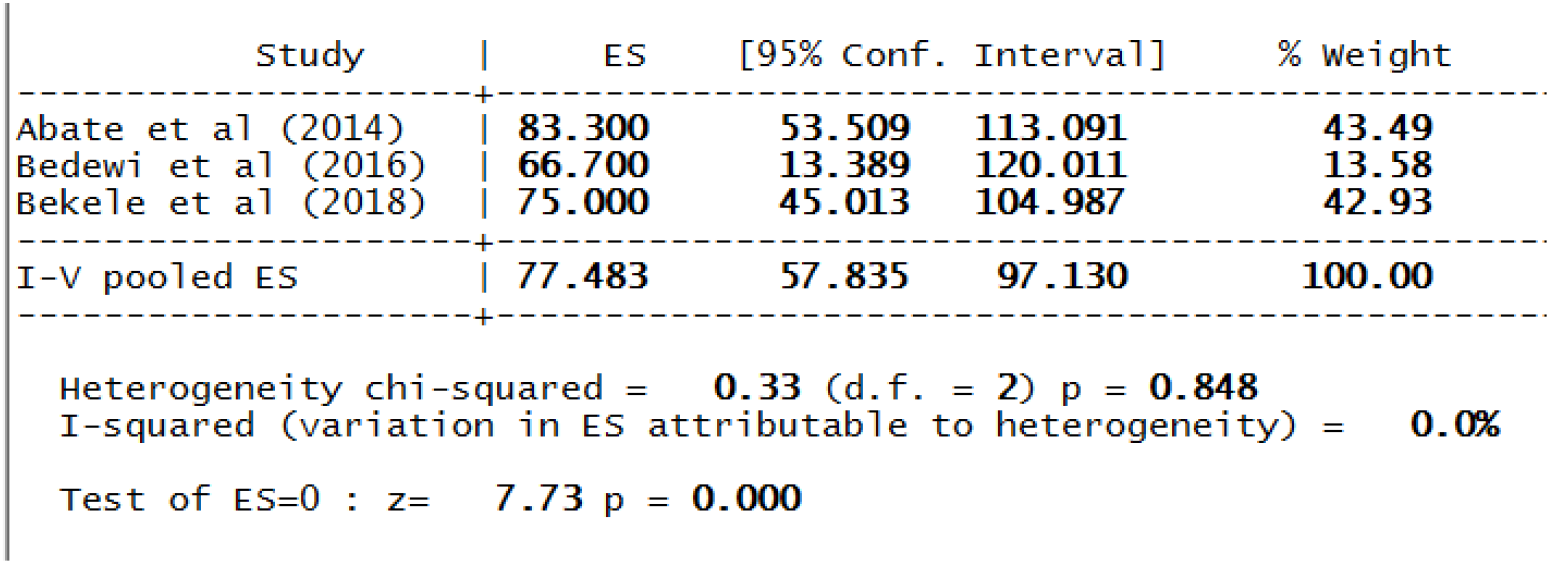
The pooled prevalence of *inh*AMUT1 (C15T) resistance among INH-resistant *Mtb* cases.

During our meta-analysis, seventeen publications evaluated 847 genotypically RIF resistant *Mtb* isolates for mutations in *rpo*B gene inclusive of an 81-base pair *β*-subunit ranged from codons 507-533, particularly, at codon 526, 526, and 531. Thus, the highest pooled estimated prevalence of the gene mutation associated with *rpo*B gene was observed in *rpo*BMUT3 (S531L), with 74.20 % (95% CI 66.39-82.00%) and the I^2^-value of 64.6% and p=0.002 (Fig 6 & Table 3). The publication bias was evaluated by using the Egger’s regression test revealed the p-value was 0.968 and a funnel plot showed a symmetrical distribution, which indicated the absence of publication bias (Fig.7). Moreover, the pooled prevalence of *rpo*BMUT2A (H526Y) was 17.20% (95% CI 8.25-26.15%) with I^2^-value of 85.7% and p<0.001 (Fig 8 & Table 3). During evaluation of publication bias, the funnel plot showed a symmetrical distribution; and the Egger’s test p-value was 0.107, which indicated there was no publication bias (Fig. 9). The analysis derived a pooled *rpo*BMUT2B(H526D) and *rpo*BMUT1(D516V) resistant *Mtb* prevalence of 13.91% and 2.96%, respectively. The absence of band at different *rpo*B gene wild type was assessed and the high pooled estimated pooled prevalence was observed in *rpo*BWT8 and *rpo*BWT3 with 58.21% and 19.92%, respectively (Table 3).

**Table 3:** Results of mutations pattern of 847 any RIF resistance *tuberculosis* patients.

| Mutation(s) |  | Frequency |  | I<sup>2</sup>(P-value) |
| --- | --- | --- | --- | --- |
| *rpoB* |  | No of patients | % (95% CI) |  |
| *rpoB*WT1 absent |  | - | - | - |
| *rpoB*WT2 - |  | 688 | 4.12%(-1.24-9.48) | 85.2%(0.001) |
| *rpoB*WT3 - |  | 645 | 19.92 (-3.37-43.21) | 93.1%(<0.001) |
| *rpoB*WT4 - |  | 688 | 9.20(0.87-17.54) | 88.1%(<0.001) |
| *rpoB*WT5 |  | - | - | - |
| *rpoB*WT6 - |  | 485 | 0.41%(-0.17-1.00) | 0.0%(0.328) |
| *rpoB*WT7 - |  | 799 | 19.30(8.78-29.82) | 90.5%(<0.001) |
| *rpoB*WT8 - |  | 806 | 58.21(26.38-90.04) | 99.1% (<0.001) |
| *rpoB*MUT1(D516V) |  | 538 | 2.96 (1.53-4.39) | 0.0% (0.503) |
| *rpoB*MUT2A (H526Y) |  | 776 | 17.20 (8.25-26.15) | 85.7% (<0.001) |
| *rpoB*MUT2B (H526D) |  | 724 | 13.91(5.80-22.02) | 87.5% (<0.001) |
| *rpoB*MUT3 (S531L) |  | 780 | 74.20(66.39-82.00) | 64.6% (0.002) |

**Figure 6:**
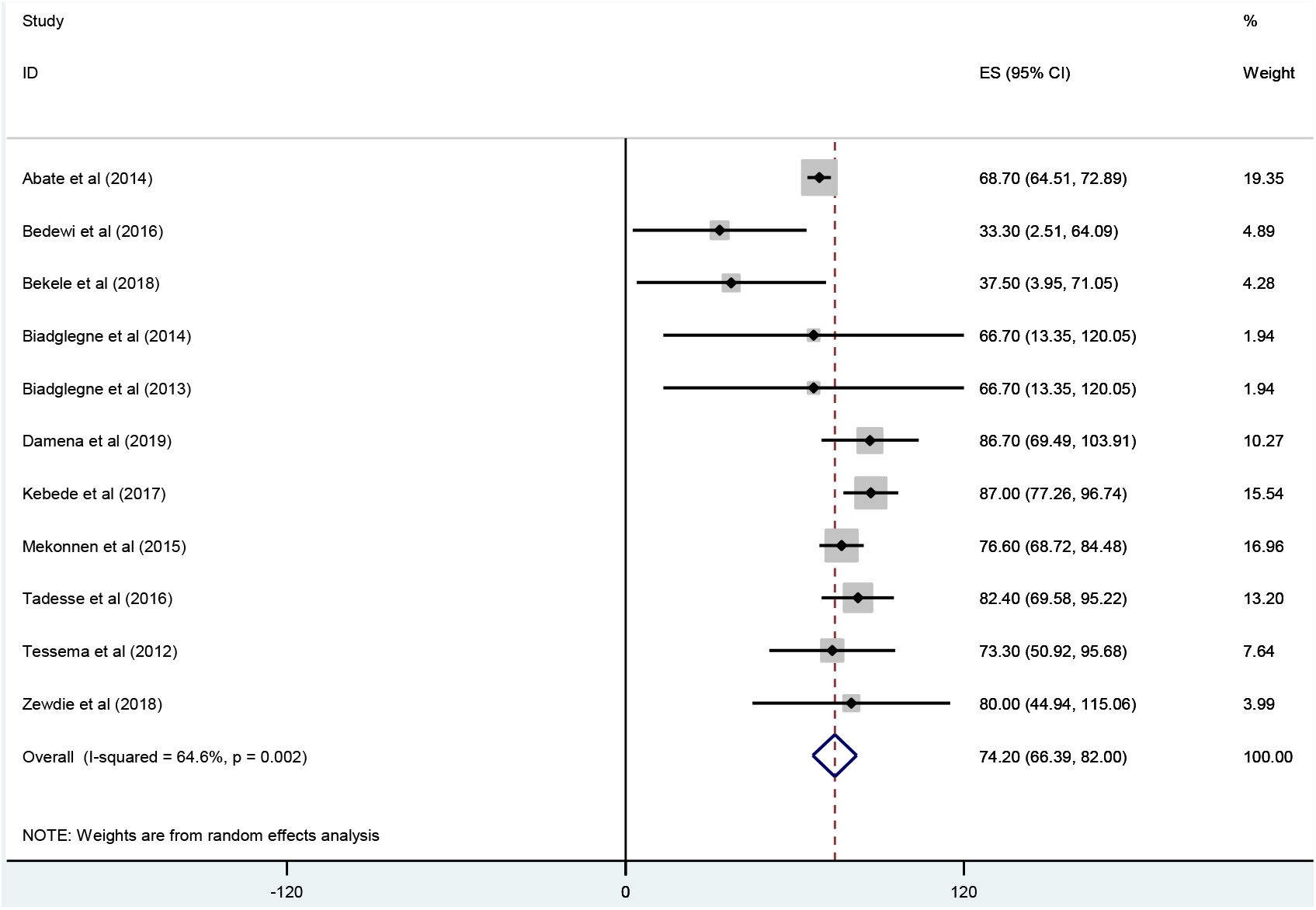
Forest plot showing the pooled prevalence of *rpo*BMUT3 (S531L) resistance among RIF-resistant *Mtb* cases.

**Figure 7:**
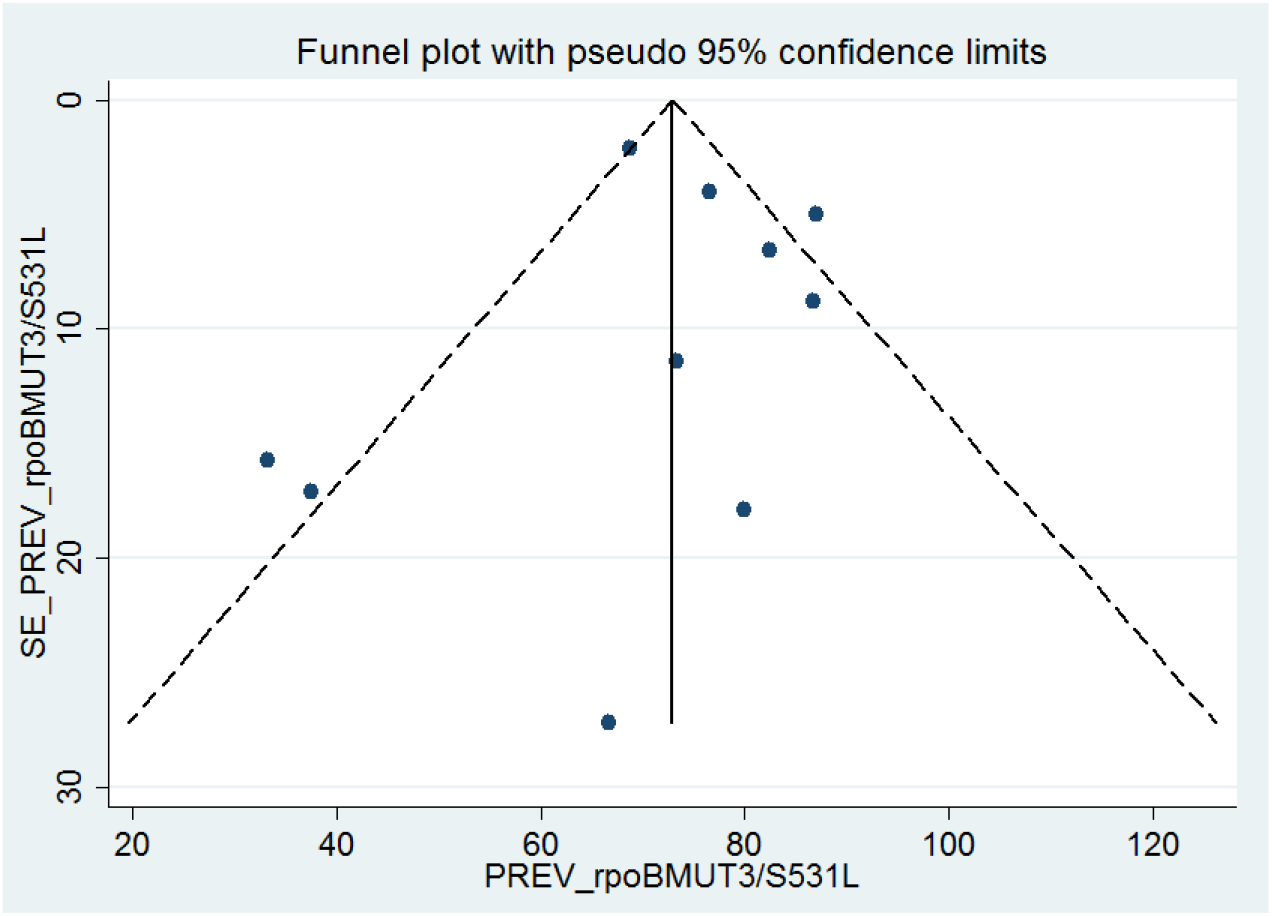
Funnel plot for publication bias, PREV (prevalence) represented in the x-axis and standard error (SE) of prevalence of *rpo*BMUT3 (S531L) in the y-axis.

**Figure 8:**
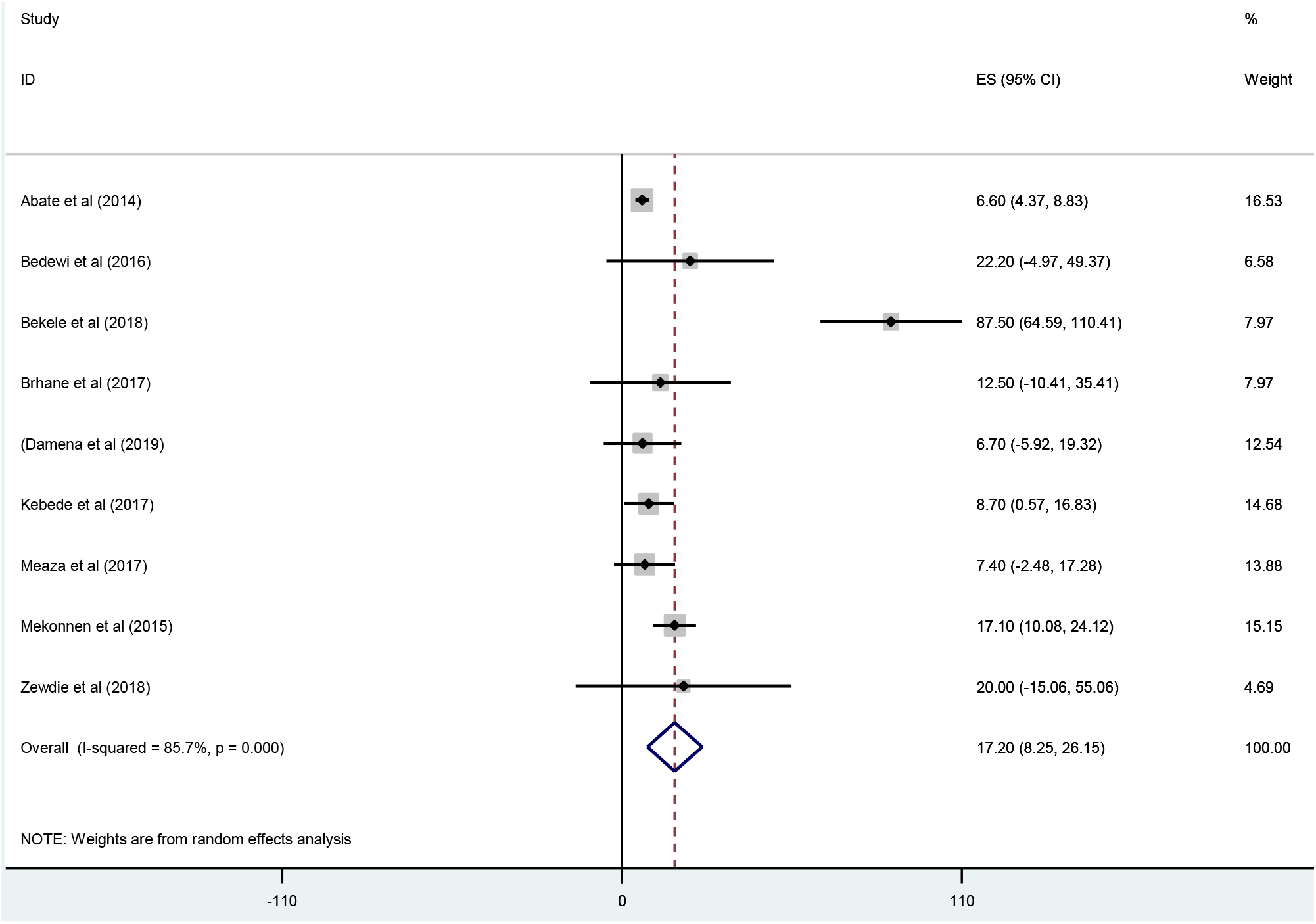
Forest plot showing the pooled prevalence of *rpo*BMUT2A (H526Y) resistance among RIF-resistant *Mtb* cases.

**Figure 9:**
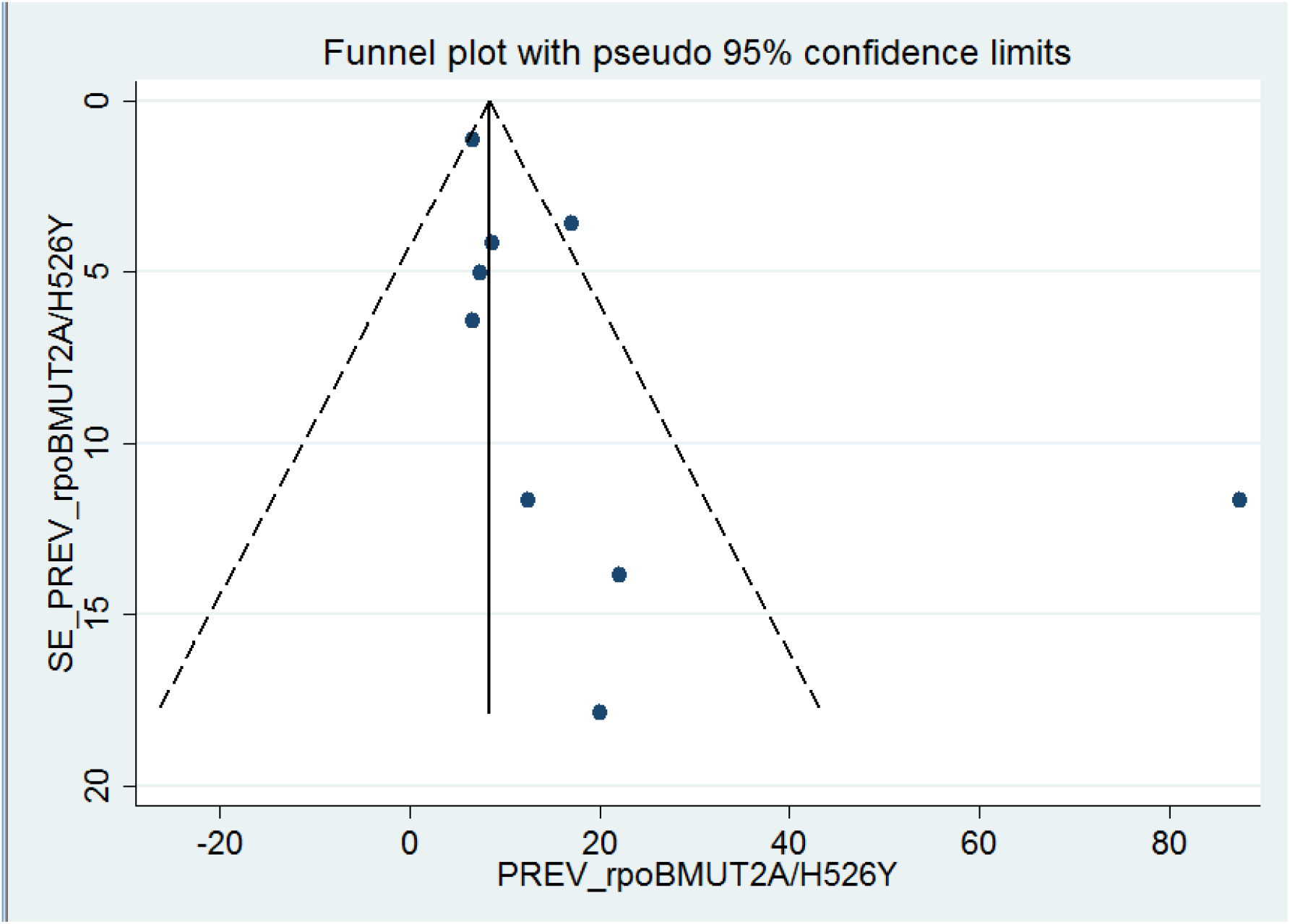
Funnel plot for publication bias, PREV (prevalence) represented in the x-axis and standard error (SE) of prevalence of *rpo*BMUT2A (H526Y) in the y-axis

## Discussion

The global TB control and prevention program is challenged due to the emergence of drug resistant bacilli [1, 2, 49], particularly due to the evolution of MDR-TB, resistance to both RIF and INH, the two “backbone” of the present recommended TB treatment [1, 3].

Isoniazid is a prodrug need to be converted into the active form by catalase-peroxidase enzyme encoded by the *kat*G gene. After activation, “INH has an effect/inhibits the bacilli’s mycolic acid synthesis through the NADH-dependent enoyl-acyl carrier protein reductase, encoded by the *inh*A gene” [50, 51]. The molecular basis of INH resistance is arbitrated by mutations in the *kat*G gene and *inh*A promotor region. The most frequent INH resistance mechanism has been identified as the *kat*GS315T gene mutation, which results an inefficient INH-NAD product inhibiting the antimicrobial actions of INH. This mechanism is associated with high-level INH resistance in MDR-TB isolates. Besides, the gene mutation at the promotor region of *inh*A gene, the most frequent occurring at the position -15, result in an overexpression of *inh*A [52-55]. However, the frequency patterns of the most common gene mutations conferring INH resistance differ between individual genes. Recent studies revealed that, the majority (64%) of INH resistance in *Mtb* isolates is associated with a single gene mutation in *kat*G315 [3, 52-55]. “The dominance of this mutation is hypothesized to be the result of a low or zero fitness cost for this mutation, allowing it to propagate without negative selection pressure” [3]. Other mutations in the *kat*G gene, other than *kat*G315 appear to occur at low (<1%) frequencies and devastatingly in combination with the *kat*G315 gene mutation [3]. This systematic review and meta-analysis demonstrated that a prevalence of 95.8% for *kat*G315 mutation and 5.9% for *inh*A promotor region gene mutation in patients with drug resistant *Mtb*, suggesting a major epidemic of DR-TB among these patients. Similarly, a study conducted by Campbell *et al*. in 212 INH-resistance in *Mtb* isolates estimated the global frequency of the *kat*G315 gene mutation to be 85%, and 17% for *inh*A-15, while the cumulative frequency was 91% [56]. A recent study revealed that the estimated global frequencies of *kat*G315 and inhA-15 was suggestively higher at 86% and 34, respectively [57]. In contrast, a meta-analysis conducted by Alagappan *et al*. revealed that the *kat*G315 mutation was 71.0%, and 29.0% for *inh*A mutation [7]. This slight inconsistency may be due to the type of TB patients and kind of INH resistance, that Alagappan *et al* estimated these gene mutations among INH-monoresistance and PTB patients, while our review assessed the prevalence of these gene mutations among any INH resistance and all type of TB patients. However, a study carried out from California (USA), identified a worldwide frequency of 61% for *kat*G315 mutation, and 23% for *inh*AMUT1(-15) mutation resembling the frequencies of these gene mutations as measured in this review [3].

Several previous studies have reported vastly variable frequencies of these mutations; with *kat*G315 gene mutations accounting for 42 to 95% and mutation in the inhA-15 promotor region accounting for 6 to 43% of phenotypic INH resistance [12, 16, 17, 22, 23].

This meta-analysis resulting a pooled *kat*GMUT1(S315T1) resistant *Mtb* prevalence of 89.18% (95% CI 81.94-96.43%), and prevalence of 0.91% (95% CI 0.195-1.63%) for *kat*GMUT2 (S315T2) resistance gene. Besides, our review estimated the prevalence of *inh*AMUT1 (C15T) at *inh*A promotor region was 77.48% (95% CI 57.84-97.13%). Similarly, Alagappan *et al*. in 1150 INH-monoresistance *Mtb* isolates from India estimated the pooled prevalence of *kat*GMUT1(S315T1) was 63.2%, while the prevalence of *kat*GMUT2(S315T2) was 0.3% [7]. This analysis revealed a robust association between the percentage of INH resistance-conferring mutation due to *kat*G (S315T) evaluated in clinical isolates and many different indicators of TB transmission intensity, supporting the suggestion that gene mutation at the 315-codon position of *kat*G confer INH resistance in *Mtb* without reducing virulence or transmissibility.

Rifampicin is one of the utmost effective antitubercular drugs, since it has significant effect against actively metabolizing and slow metabolizing bacilli, making RIF a key component of the existing first line treatment option used to the treatment of drug sensitive TB (DS-TB) [16, 50, 51, 58]. The gene mutations associated with RIF resistance are well known and seem to occur in a single gene region of phenotypically RIF resistant *Mtb* isolates [7].

In *Mtb*, RIF binds to the *β*-subunit of the RNA polymerase, subsequently inhibiting the elongation of mRNA. RIF resistance in *Mtb* is mediated by the gene mutations clustered in codons 507-533 of the gene coding for the RNA polymerase *β*-subunit (*rpo*B) [49, 50].

Although, the gene mutations outside the RIF resistance-determining region (RRDR) have been reported in RIF resistant *Mtb* isolates [59], the 81-base pair *β*-subunit (*rpo*B) region is known as RRDR, which is the target of current genotypic/molecular based assays and accounts for 96% of RIF resistance. The amino acid or codons 531 and 526 comprise the most frequent gene mutations associated with RIF resistance in *Mtb* isolates [50, 51, 60-64]. RIF monoresistance is rare and it occurs in combination with resistance to other drugs, most frequently INH, making RIF targets a surrogate marker of the MDR-TB phenotype [65]. Several studies revealed that RIF resistance commonly occurs due to *rpo*B gene mutation, which accounts approximately 96% of RIF resistance [50, 51, 60-64]. This review demonstrated that a prevalence of 90.8% for *rpo*B gene mutation in patients with DR-TB, suggesting a serious epidemic of RIF resistant *Mtb* among these patients. The present meta-analysis derived the most common gene mutation associated with RIF resistance was *rpo*BMUT3(S531L) with a pooled prevalence of 74.20 % (95% CI 66.39-82.00%). Besides, the second most frequent RIF resistance conferring mutation in this review was *rpo*BMUT2A (H526Y) with 17.20% estimated pooled prevalence. Similarly, a study conducted by Elbir *et al*. from Sudan reported that the frequency of gene mutation at codon 531, 526 and 516 were 64.1%, 17.9%, and 7.7%, respectively [9]. The same repot revealed that all genetic alterations were occurred by single base substitutions, and the most common mutation was observed at codon S531L [9]. Several previous studies have reported supporting evidence to our review result, that the most common gene mutation associated with RIF resistance in *Mtb* is due to the nucleotide (codon) change at 531 and 526 in *rpo*B/RRDR [6, 9, 49, 66].

## Limitations

As a limitation, first, in this systematic review and meta-analysis, only English published articles were included for the analysis. Second, due to lack of detail information in few included studies, this review did not present prevalence of RIF and INH monoresistance and have not estimated the pooled prevalence of gene mutations associated with RIF and INH monoresistance. Third, majority of the studies have not explained proportions of gene mutations based on sex and age of participants; so that, this review did not explain sex and age-wise comparison of RIF and INH resistance mutations. Four, the development of gene mutations in *Mtb* strains associated with different anti-TB drug resistance varies across different TB patient’s treatment outcome (failure, loss of follow-up, retreatment cases) and diagnostic test results (e.g. retreatment smear negative and positive case, any follow-up smear positive cases) as well as other patient’s associated health conditions; however, due to the lack of detail information regarding these issues, this review failed to explain the estimated pooled prevalence of different gene mutations associated with RIF and INH among those TB patient categories.

## Conclusions

In conclusion, RIF resistance was most commonly occurred due to mutations in the S531L followed by H526Y genes, while INH resistance was most frequently due to mutations in the *kat*G315 gene, and these mutations were also associated with MDR and polydrug resistance, whereas mutations in *inh*A promotor region were less frequent. Commonly, the gene mutations in both *kat*G gene and *inh*A promotor region rise the development of MDR-TB and the risk of relapse. However, the increasing frequencies of these gene mutations seem to vary by region, which could lead to differences in the sensitivity of genotypic/molecular diagnostics tools, if the tests are based only on these gene mutations. This would permit for modifying of genotypic/molecular tests to specific geographical locations, better understanding/interpretation of the molecular tests being used, and better therapy recommendations.

## Data Availability

The datasets anlyzed during this review can be accessed from the corrosponding author upon reasonable request.

## Abbreviations

DR-TB: drug resistant tuberculosis
DST: drug susceptibility testing
DS-TB: drug senstetive tuberculosis
INH: isoniazid
MDR-TB: multidrug resistant tuberculosis
*Mtb*: *Mycobacterium tuberculosis*
PRISMA: Preferred Reporting Items for Systematic review and Meta-analysis
RIF: rifampicin
TB: tuberculosis
WHO: world health organization
XDR-TB: extensive drug resistance TB.

## Authors’ contributions

MAR: conceived the study, took the primary role in data acquisition, formal analysis, interpretation, writing the manuscrpit for publication. BA and BBA: took revision of the study protocol, particpated in result interpretation and review final manuscrpit for publication. All authors read and approved the final maunscrpit for publication.

## Transparency declaration

The authors declare no conflict of interest

## Acknowledgments

Our aknowldgement goes to all corrosponding authors who provided us information when we requested them.

---

**Supplementary Figure S1**. Representative DNA patterns obtained by the GenoType®MTBDR*plus* assay.

**Supplementary Table S1:** The PRISMA checklist.

**Supplementary Table S2**: Table S2. Search strategy used for one of the databases (Medline/PubMed)

**Supplementary Table S3:** Quality appraisal result of included studies; Using Joanna Briggs Institute (JBI) quality appraisal checklist for cross-sectional studies.

**Supplementary Table S4:** Characteristics of included studies.

**Supplementary Table S5:** Characteristics of included studies.

## References

1. Global tuberculosis report 2019. Geneva. World Health Organization; 2019. Licence: CC BY-NC-SA 3.0 IGO. 2019.

2. Abebe G, Paasch F, Apers L, Rigouts L, Colebunders R. Tuberculosis drug resistance testing by molecular methods: opportunities and challenges in resource limited settings. J Microbiol Methods. 2011;84(2):155–60.

3. Seifert M, Catanzaro D, Catanzaro A, Rodwell TC. Genetic mutations associated with isoniazid resistance in Mycobacterium tuberculosis: a systematic review. PloS one. 2015;10(3).

4. Marahatta SB, Gautam S, Dhital S, Pote N, Jha AK, Mahato R, et al. katG (SER 315 THR) gene mutation in isoniazid resistant Mycobacterium tuberculosis. Kathmandu Univ Med J (KUMJ). 2011;9(33):19–23.

5. Rueda J, Realpe T, Mejia GI, Zapata E, Rozo JC, Ferro BE, et al. Genotypic Analysis of Genes Associated with Independent Resistance and Cross-Resistance to Isoniazid and Ethionamide in Mycobacterium tuberculosis Clinical Isolates. Antimicrob Agents Chemother. 2015;59(12):7805–10.

6. Kigozi E, Kasule GW, Musisi K, Lukoye D, Kyobe S, Katabazi FA, et al. Prevalence and patterns of rifampicin and isoniazid resistance conferring mutations in Mycobacterium tuberculosis isolates from Uganda. PLoS One. 2018;13(5):e0198091.

7. Alagappan C, Shivekar SS, Brammacharry U, Kapalamurthy VRC, Sakkaravarthy A, Subashkumar R, et al. Prevalence of mutations in genes associated with isoniazid resistance in Mycobacterium tuberculosis isolates from re-treated smear-positive pulmonary tuberculosis patients: A meta-analysis. Journal of global antimicrobial resistance. 2018;14:253–9.

8. Nguyen L. Antibiotic resistance mechanisms in M. tuberculosis: an update. Arch Toxicol. 2016;90(7):1585–604.

9. Elbir H, Ibrahim NY. Frequency of mutations in the rpoB gene of multidrug-resistant Mycobacterium tuberculosis clinical isolates from Sudan. J Infect Dev Ctries. 2014;8(6):796–8.

10. Bollela VR, Namburete EI, Feliciano CS, Macheque D, Harrison LH, Caminero JA. Detection of katG and inhA mutations to guide isoniazid and ethionamide use for drug-resistant tuberculosis. Int J Tuberc Lung Dis. 2016;20(8):1099–104.

11. Lessells RJ, Cooke GS, Newell M-L, Godfrey-Faussett P. Evaluation of tuberculosis diagnostics: establishing an evidence base around the public health impact. Journal of Infectious Diseases. 2011;204(suppl_4):S1187-S95.

12. Zhang Y, Yew W. Mechanisms of drug resistance in Mycobacterium tuberculosis [State of the art series. Drug-resistant tuberculosis. Edited by CY. Chiang. Number 1 in the series]. The International Journal of Tuberculosis and Lung Disease. 2009;13(11):1320–30.

13. Scior T, Meneses Morales I, Garcés Eisele SJ, Domeyer D, Laufer S. Antitubercular isoniazid and drug resistance of Mycobacterium tuberculosis—a review. Archiv der Pharmazie: An International Journal Pharmaceutical and Medicinal Chemistry. 2002;335(11-12):511–25.

14. Laurenzo D, Mousa SA. Mechanisms of drug resistance in Mycobacterium tuberculosis and current status of rapid molecular diagnostic testing. Acta Trop. 2011;119(1):5–10.

15. Riska PF, Jacobs JW, Alland D. Molecular determinants of drug resistance in tuberculosis. The International Journal of Tuberculosis and Lung Disease. 2000;4 (2 Suppl 1):S4–10.

16. Rattan A, Kalia A, Ahmad N. Multidrug-resistant Mycobacterium tuberculosis: molecular perspectives. Emerging infectious diseases. 1998;4(2):195.

17. Rossetti M, Valim A, Silva M, Rodrigues V. Resistant tuberculosis: a molecular review. Rev Saude Publica. 2002;36(4):525–32.

18. Zhang Y, Heym B, Allen B, Young D, Cole S. The catalase—peroxidase gene and isoniazid resistance of Mycobacterium tuberculosis. Nature. 1992;358(6387):591–3.

19. Vilchèze C, Wang F, Arai M, Hazbón MH, Colangeli R, Kremer L, et al. Transfer of a point mutation in Mycobacterium tuberculosis inhA resolves the target of isoniazid. Nature medicine. 2006;12(9):1027–9.

20. Telenti A, Imboden P, Marchesi F, Matter L, Schopfer K, Bodmer T, et al. Detection of rifampicin-resistance mutations in Mycobacterium tuberculosis. The Lancet. 1993;341(8846):647–51.

21. Sekyere JO, Reta MA, Maningi NE, Fourie PB. Antibiotic resistance of Mycobacterium tuberculosis complex in Africa: A systematic review of current reports of molecular epidemiology, mechanisms and diagnostics. J Infect. 2019;79(6):550–71.

22. Ahmad S, Mokaddas E. Recent advances in the diagnosis and treatment of multidrug-resistant tuberculosis. Respir Med CME. 2010;3(2):51–61.

23. Sreevatsan S, Pan X, Zhang Y, Deretic V, Musser JM. Analysis of the oxyR-ahpC region in isoniazid-resistant and-susceptible Mycobacterium tuberculosis complex organisms recovered from diseased humans and animals in diverse localities. Antimicrob Agents Chemother. 1997;41(3):600–6.

24. Kwak N, Choi SM, Lee J, Park YS, Lee C-H, Lee S-M, et al. Diagnostic accuracy and turnaround time of the Xpert MTB/RIF assay in routine clinical practice. PloS one. 2013;8(10).

25. Yadav RN, Singh BK, Sharma SK, Sharma R, Soneja M, Sreenivas V, et al. Comparative evaluation of GenoType MTBDRplus line probe assay with solid culture method in early diagnosis of multidrug resistant tuberculosis (MDR-TB) at a tertiary care centre in India. PloS one. 2013;8(9).

26. Abate G, Hoffner S, Thomsen VØ, Miörner H. Characterization of isoniazid-resistant strains of Mycobacterium tuberculosis on the basis of phenotypic properties and mutations in katG. Eur J Clin Microbiol Infect Dis. 2001;20(5):329–33.

27. Moher D, Liberati A, Tetzlaff J, Altman DG, Group P. Preferred reporting items for systematic reviews and meta-analyses: the PRISMA statement. PLoS Med. 2009;6(7):e1000097.

28. Joanna Briggs Institute. The Joanna Briggs Institute Critical Appraisal tools for use in JBI Systematic Reviews: Checklist for Prevalence Studies. Retreived November. 2017;15:2018.

29. Egger M, Smith GD, Schneider M, Minder C. Bias in meta-analysis detected by a simple, graphical test. Bmj. 1997;315(7109):629–34.

30. Abate D, Tedla Y, Meressa D, Ameni G. Isoniazid and rifampicin resistance mutations and their effect on second-line anti-tuberculosis treatment. Int J Tuberc Lung Dis. 2014;18(8):946–51.

31. Amir Alelign, Aboma Zewude, Temesgen Mohammed, Samuel Tolosa, Gobena Ameni, Beyene Petros. Molecular detection of Mycobacterium tuberculosis sensitivity to rifampicin and isoniazid in South Gondar Zone, northwest Ethiopia. BMC Infectious Diseases. 2019;19:343.

32. Bedewi Omer Z, Mekonnen Y, Worku A, Zewde A, Medhin G, Mohammed T, et al. Evaluation of the GenoType MTBDRplus assay for detection of rifampicin- and isoniazid-resistant Mycobacterium tuberculosis isolates in central Ethiopia. Int J Mycobacteriol. 2016;5(4):475–81.

33. Bekele S, Derese Y, Hailu E, Mihret A, Dagne K, Yamuah L, et al. Line-probe assay and molecular typing reveal a potential drug resistant clone of Mycobacterium tuberculosis in Ethiopia. Tropical diseases, travel medicine and vaccines. 2018;4:15.

34. Beyene D, Bergval I, Hailu E, Ashenafi S, Yamuah L, Aseffa A, et al. Identification and genotyping of the etiological agent of tuberculous lymphadenitis in Ethiopia. J Infect Dev Ctries. 2009;3(6):412–9.

35. Biadglegne F, Mulu A, Rodloff AC, Sack U. Diagnostic performance of the Xpert MTB/RIF assay for tuberculous lymphadenitis on fine needle aspirates from Ethiopia. Tuberculosis (Edinb). 2014;94(5):502–5.

36. Biadglegne F, Tessema B, Rodloff AC, Sack U. Magnitude of gene mutations conferring drug resistance in mycobacterium tuberculosis isolates from lymph node aspirates in ethiopia. Int J Med Sci. 2013;10(11):1589–94.

37. Brhane M, Kebede A, Petros Y. Molecular detection of multidrug-resistant tuberculosis among smear-positive pulmonary tuberculosis patients in Jigjiga town, Ethiopia. Infection and drug resistance. 2017;10:75.

38. Damena D, Tolosa S, Hailemariam M, Zewude A, Worku A, Mekonnen B, et al. Genetic diversity and drug susceptibility profiles of Mycobacterium tuberculosis obtained from Saint Peter’s TB specialized Hospital, Ethiopia. PLoS One. 2019;14(6):e0218545.

39. Haile B, Tafess K, Zewude A, Yenew B, Siu G, Ameni G. Spoligotyping and drug sensitivity of Mycobacterium tuberculosis isolated from pulmonary tuberculosis patients in the Arsi Zone of southeastern Ethiopia. New Microbes New Infect. 2020;33:100620.

40. Kebede A, Demisse D, Assefa M, Getachew Z, Yenew B, Tedla Y, et al. Performance of MTBDRplus assay in detecting multidrug resistant tuberculosis at hospital level. BMC Res Notes. 2017;10(1):661.

41. Meaza A, Kebede A, Yaregal Z, Dagne Z, Moga S, Yenew B, et al. Evaluation of genotype MTBDRplus VER 2.0 line probe assay for the detection of MDR-TB in smear positive and negative sputum samples. BMC Infect Dis. 2017;17(1):280.

42. Mekonnen D, Admassu A, Mulu W, Amor A, Benito A, Gelaye W, et al. Multidrug-resistant and heteroresistant Mycobacterium tuberculosis and associated gene mutations in Ethiopia. Int J Infect Dis. 2015;39:34–8.

43. Tadesse M, Abebe G, Bekele A, Bezabih M, de Rijk P, Meehan CJ, et al. The predominance of Ethiopian specific Mycobacterium tuberculosis families and minimal contribution of Mycobacterium bovis in tuberculous lymphadenitis patients in Southwest Ethiopia. Infection, Genetics and Evolution. 2017;55:251–9.

44. Tadesse M, Aragaw D, Dimah B, Efa F, Abdella K, Kebede W, et al. Drug resistance-conferring mutations in Mycobacterium tuberculosis from pulmonary tuberculosis patients in Southwest Ethiopia. Int J Mycobacteriol. 2016;5(2):185–91.

45. Tessema B, Beer J, Emmrich F, Sack U, Rodloff AC. Analysis of gene mutations associated with isoniazid, rifampicin and ethambutol resistance among Mycobacterium tuberculosis isolates from Ethiopia. BMC Infect Dis. 2012;12:37.

46. Wondale B, Medhin G, Abebe G, Tolosa S, Mohammed T, Teklu T, et al. Phenotypic and genotypic drug sensitivity of Mycobacterium tuberculosis complex isolated from South Omo Zone, Southern Ethiopia. Infect Drug Resist. 2018;11:1581–9.

47. Workalemahu B, Berg S, Tsegaye W, Abdissa A, Girma T, Abebe M, et al. Genotype diversity of Mycobacterium isolates from children in Jimma, Ethiopia. BMC Res Notes. 2013;6:352.

48. Zewdie O, Mihret A, Abebe T, Kebede A, Desta K, Worku A, et al. Genotyping and molecular detection of multidrug-resistant Mycobacterium tuberculosis among tuberculosis lymphadenitis cases in Addis Ababa, Ethiopia. New Microbes and New Infections. 2018;21:36–41.

49. Dookie N, Rambaran S, Padayatchi N, Mahomed S, Naidoo K. Evolution of drug resistance in Mycobacterium tuberculosis: a review on the molecular determinants of resistance and implications for personalized care. J Antimicrob Chemother. 2018;73(5):1138–51.

50. Palomino JC, Martin A. Drug Resistance Mechanisms in Mycobacterium tuberculosis. Antibiotics (Basel, Switzerland). 2014;3(3):317–40.

51. Almeida Da Silva PE, Palomino JC. Molecular basis and mechanisms of drug resistance in Mycobacterium tuberculosis: classical and new drugs. Journal of antimicrobial chemotherapy. 2011;66(7):1417–30.

52. Ramaswamy SV, Reich R, Dou S-J, Jasperse L, Pan X, Wanger A, et al. Single nucleotide polymorphisms in genes associated with isoniazid resistance in Mycobacterium tuberculosis. Antimicrob Agents Chemother. 2003;47(4):1241–50.

53. Hazbón MH, Brimacombe M, Del Valle MB, Cavatore M, Guerrero MI, Varma-Basil M, et al. Population genetics study of isoniazid resistance mutations and evolution of multidrug-resistant Mycobacterium tuberculosis. Antimicrob Agents Chemother. 2006;50(8):2640–9.

54. Vilchèze C, Jacobs J, William R. The mechanism of isoniazid killing: clarity through the scope of genetics. Annu Rev Microbiol. 2007;61:35–50.

55. Banerjee A, Dubnau E, Quemard A, Balasubramanian V, Um KS, Wilson T, et al. inhA, a gene encoding a target for isoniazid and ethionamide in Mycobacterium tuberculosis. Science. 1994;263(5144):227–30.

56. Campbell PJ, Morlock GP, Sikes RD, Dalton TL, Metchock B, Starks AM, et al. Molecular detection of mutations associated with first-and second-line drug resistance compared with conventional drug susceptibility testing of Mycobacterium tuberculosis. Antimicrob Agents Chemother. 2011;55(5):2032–41.

57. Rodwell TC, Valafar F, Douglas J, Qian L, Garfein RS, Chawla A, et al. Predicting extensively drugresistant Mycobacterium tuberculosis phenotypes with genetic mutations. Journal of clinical microbiology. 2014;52(3):781–9.

58. Mitchison DA. Basic mechanisms of chemotherapy. Chest. 1979;76(6):771–80.

59. Heep M, Rieger U, Beck D, Lehn N. Mutations in the beginning of the rpoBGene can induce resistance to Rifamycins in both helicobacter pylori and mycobacterium tuberculosis. Antimicrob Agents Chemother. 2000;44(4):1075–7.

60. Palomino JC, Martin A. Drug resistance mechanisms in Mycobacterium tuberculosis. Antibiotics. 2014;3(3):317–40.

61. Blanchard JS. Molecular mechanisms of drug resistance in Mycobacterium tuberculosis. Annu Rev Biochem. 1996;65(1):215–39.

62. Telenti A, Imboden P, Marchesi F, Schmidheini T, Bodmer T. Direct, automated detection of rifampin-resistant Mycobacterium tuberculosis by polymerase chain reaction and single-strand conformation polymorphism analysis. Antimicrob Agents Chemother. 1993;37(10):2054–8.

63. Somoskovi A, Parsons LM, Salfinger M. The molecular basis of resistance to isoniazid, rifampin, and pyrazinamide in Mycobacterium tuberculosis. Respir Res. 2001;2(3):164.

64. Caws M, Duy PM, Tho DQ, Lan NTN, Farrar J. Mutations prevalent among rifampin-and isoniazid-resistant Mycobacterium tuberculosis isolates from a hospital in Vietnam. Journal of clinical microbiology. 2006;44(7):2333–7.

65. Traore H, Fissette K, Bastian I, Devleeschouwer M, Portaels F. Detection of rifampicin resistance in Mycobacterium tuberculosis isolates from diverse countries by a commercial line probe assay as an initial indicator of multidrug resistance. The international journal of tuberculosis and lung disease. 2000;4(5):481–4.

66. Takawira FT, Mandishora RSD, Dhlamini Z, Munemo E, Stray-Pedersen B. Mutations in rpoB and katG genes of multidrug resistant mycobacterium tuberculosis undetectable using genotyping diagnostic methods. Pan Afr Med J. 2017;27:145.

